# Predicting Alzheimer’s Disease with Multi-Omic Data: A Systematic Review

**DOI:** 10.1101/2022.11.25.22282770

**Authors:** Anthony Davis, Wilson Mendoza, Daniel Leach, Oge Marques

## Abstract

**Background and Purpose:** Alzheimer’s Disease (AD) is a complex neurodegenerative disease that has been becoming increasingly prevalent in recent decades. Efforts to identify predictive biomarkers of the disease have proven difficult. Advances in the collection of multi-omic data and deep learning algorithms have opened the possibility of integrating these various data together to identify robust biomarkers for predicting the onset of the disease prior to the onset of symptoms. This study performs a systematic review of recent methods used to predict AD using multi-omic and multi-modal data.

**Methods:** We systematically reviewed studies from Google Scholar, Pubmed, and Semantic Scholar published after 2018 in relation to predicting AD using multi-omic data. Three reviewers independently identified eligible articles and came to a consensus of papers to review. The Quality in Prognosis Studies (QUIP) tool was used for the risk of bias assessment.

**Results:** 22 studies which use multi-omic data to either predict AD or develop AD biomarkers were identified. Those studies which aimed to directly classify AD or predict the progression of AD achieved area under the receiver operating characteristic curve (AUC) between .70 - .98 using varying types of patient data, most commonly extracted from blood. Hundreds of new genes, single nucleotide polymorphisms (SNPs), RNA molecules, DNA methylation sites, proteins, metabolites, lipids, imaging features, and clinical data have been identified as successful biomarkers of AD. The most successful techniques to predict AD have integrated multi-omic data together in a single analysis.

**Conclusion:** This review has identified many successful biomarkers and biosignatures that are less invasive than cerebral spinal fluid. Together with the appropriate prediction models, highly accurate classifications and prognostications can be made for those who are at risk of developing AD. These early detection of risk factors may help prevent the further development of cognitive impairment and improve patient outcomes.

## Introduction

Alzheimer’s Disease (AD) is a form of dementia generally seen in advanced age which results in the gradual decline of cognitive function. 10 % of individuals over the age of 65 have AD [1]. Cases of AD have been rising due to our aging population and are expected to triple by 2050 [2]. This will incur a high societal and economic cost that will hamper our healthcare systems and burden caregivers.

In order to best deal with this rising prevalence of AD, it is of the utmost importance to be able to identify signs of AD in the earliest possible stages. Much research has been conducted to investigate the mechanisms involved in AD, but the full pathophysiology has not yet been fully explained [3]. Known physiological changes include amyloid plaque deposits and neurofibrillary tangle formation, but the pathogenesis of these effects is unknown and no effective therapies have been produced. The ineffectiveness of proposed therapies that target these changes in the brain further suggests that AD may be more complex than the accumulation of amyloid plaques, and may in fact be an autoimmune disease [4–7]. It is clear that the current state of research does not understand AD sufficiently enough to be able to treat it post-diagnosis, so the current best option for dealing with AD is prevention.

Significant progress has been made in identifying biomarkers from genomics, blood, imaging, and cerebral spinal fluid (CSF) [8], [9]. With the emergence of affordable sequencing technologies, genomic data has become increasingly abundant and has had some success in identifying AD risk factor genes. The APOE 4 allele has been identified as one of the most closely associated genetic factors with AD. However, by itself, it is not a good predictor of AD because only half of APOE 4 carriers actually develop the disease and several studies have failed to reproduce the connection between APOE 4 and AD [10], [11]. Blood panels using protein signatures have been able to produce up to 90 accuracy for classifying AD in symptomatic patients, yet these studies have struggled to reproduce consistent protein biomarkers and results [12],[13]. CSF is a more predictive source of biomarkers due to its contact with the central nervous system, however, because it is more invasive to extract, there is less data available on CSF biomarkers. Efforts to study CSF have produced biomarkers such as -amyloid 42 and tau protein with 90% accuracy in identifying symptomatic AD [14]and can predict symptoms up to 25 years before the onset of symptoms [15].

By themselves, these various biomarkers are mixed in their usefulness in providing patients and clinicians actionable information. They may help to identify individuals who are at risk to develop AD, or they may help diagnose AD, but they are not predictive enough to prescribe lifestyle changes that may prevent the development of AD before it becomes symptomatic [16],[17]. Integrating multiple biomarkers into a combined risk score has shown promise in improving the accuracy of predicting the onset of AD [8][9]. The emergence of new “omics” categories with the development of high-throughput bioinformatics tools has created more opportunities for researchers to identify noninvasive and highly predictive AD biomarkers. By combining various types of data about the body, a more mechanistic understanding of disease can be formed and better predictions of disease progression can be made.

At the same time, deep learning has caused a paradigm shift in how big data is processed. Deep learning techniques are especially well suited to integrating multiple modes of data together to expose patterns that would otherwise be difficult for a human to identify. Using integrated multi-omic data with modern deep learning techniques shows great promise in identifying less invasive and more predictive biomarkers in AD and other diseases. A similar systematic review was conducted by Chen et al. in 2021 [18] to review predictive models for conversion of Mild Cognitive Impairment (MCI) to AD. Because this review was constrained to studies involving patients with symptomatic MCI, we consider this review necessary to discuss the effectiveness of predicting AD prior to the onset of MCI.

### Objectives

Our objectives are to identify the types of omic data and types of data processing techniques which can best predict the onset of AD. Our key questions are:

Which combinations of omics data produce the most predictive biomarkers for Alzheimer’s disease? What data processing techniques are best at deriving these biomarkers? What are the measures of accuracy for predicting the onset of AD given these biomarkers, and how early can an accurate prediction be made?

## Methods

The databases searched in this review were Google Scholar, Pubmed, and Semantic Scholar. The search terms used were specific to each database due to the difference in search operators between the databases. Of particular interest to this review is the use of machine learning or artificial intelligence (AI) in identifying biomarkers, so these words are included in our search terms. The following is a list of the search terms and operators used for each database:

### Pubmed

1. Artificial Intelligence omics predict Alzheimer’s
2. omics Alzheimer’s prognosis
3. omics predict diagnosis Alzheimer’s
4. machine learning omics predict Alzheimer’s

### Google Scholar

1. “Artificial Intelligence” “multi omics” “predict” “Alzheimer’s” “diagnosis”
2. ”machine learning” “multi omics” “predict” “Alzheimer’s” “prognosis”
3. “multi-omics data” “Alzheimer’s disease” “prognosis prediction”

### Semantic Scholar

1. “machine learning” “omics” “Alzheimer’s” “predict prognosis”
2. “artificial intelligence” “omics” “Alzheimer’s” “predict diagnosis”

Articles published prior to 2018 were filtered out in order to focus on only the most recent developments in this space. From this search strategy, a list of 553 articles were identified.

### Eligibility Criteria

To be included in this review, the identified articles were required to meet the following criteria:

1. Must use a data driven computational technique to predict AD biomarkers or AD directlyAt least two distinct varieties of omics data must be used to make the prediction
2. The data must come from humans only
3. It must be published in English
4. It must be published later than 2018

### Criteria for an article being excluded from this review are

1. Review article or unoriginal data
2. Non-open access article or otherwise requiring a fee to view
3. No quantification of the predictiveness of the technique and no biomarkers reported
4. Inability to find the original publication in its entirety

### Selection Process

The initial list of studies was identified from the three databases using these previous discussed search terms and filtering for studies published after 2018. Duplicates and studies published in languages other than English were removed.

With this list of remaining articles and the eligibility criteria, the first three authors (A.D., W.M., and D.L.) would separately and independently rate each article to be included, excluded, or unsure based on the title of the article. This was done using a shared spreadsheet through Google Sheets. If all the three researchers rated an article to be excluded, it was removed from the list of eligible articles.

After the initial filtering via the title was complete, each of the remaining studies’ abstract was read and a similar independent rating was conducted to include, exclude or discuss an article. The independent ratings were again conducted, and an online voice discussion was held to discuss each article for which there was any disagreement. Each article without a consensus was discussed until all three first authors agreed whether to include or exclude each article

No automation review tools used for this process, only Google Drive as a shared file storage.

Researchers W.M. and D.L. collected data independently from half of the 24 selected studies. 7 specific data items were sought in each study: the data types used in the study, the datasets used, the biomarkers identified, the sample size of the data, the source of the data, the validation scheme, the type of algorithm used to assess the data, and the predictive strength of these biomarkers. These data items were collected in a single shared spread sheet. Their collections were reviewed by the other two reviewers for accuracy and clarity. Secondary information collected was the country the study was primarily conducted in and code availability.

The included studies used a variety of metrics for model predictive performance. Accuracy and AUROC (Area Under Receiver Operating Characteristic) curve were the most common metrics, but studies included C-index, log2FC, and CDR as their effect measures. Only AUC, C-index and accuracy were recorded for simplicity.

### Risk of Bias Assessment

Bias is assessed using the Quality in Prognosis Studies (QUIPS) tool [19]. QUIPS assesses the risk of bias in predictor finding studies, such as those to identify AD biomarkers, so it is appropriate for this review. QUIPS consists of 6 stages: study participation, study attrition, prognostic factor measurement, outcome measurement, study confounding and statistical analysis and reporting. Three researchers (A.D., D.L., and W.M.) independently rated the risk of bias in these 6 categories as low, medium, or high. For studies with greater than 25% rating disagreement, a group discussion was held until a consensus could be reached. Initial disagreement between the researchers occurred on 4 studies, Abdullah et al., Corces et al., Gupta et al., and Song et al.

## Results

### Study Selection

553 studies were identified from the initial search of the three databases. 162 of those studies were duplicates, and 4 were written in a language other than English. 387 articles were reviewed for inclusion based on their title, and 317 of them were excluded. 70 studies remained which were reviewed using their abstract. 48 articles were removed based on the eligibility criteria after the abstract was considered, and a final set of 22 articles were identified to be included. The results of this selection process can be seen in figure 1.

**Figure 1:**
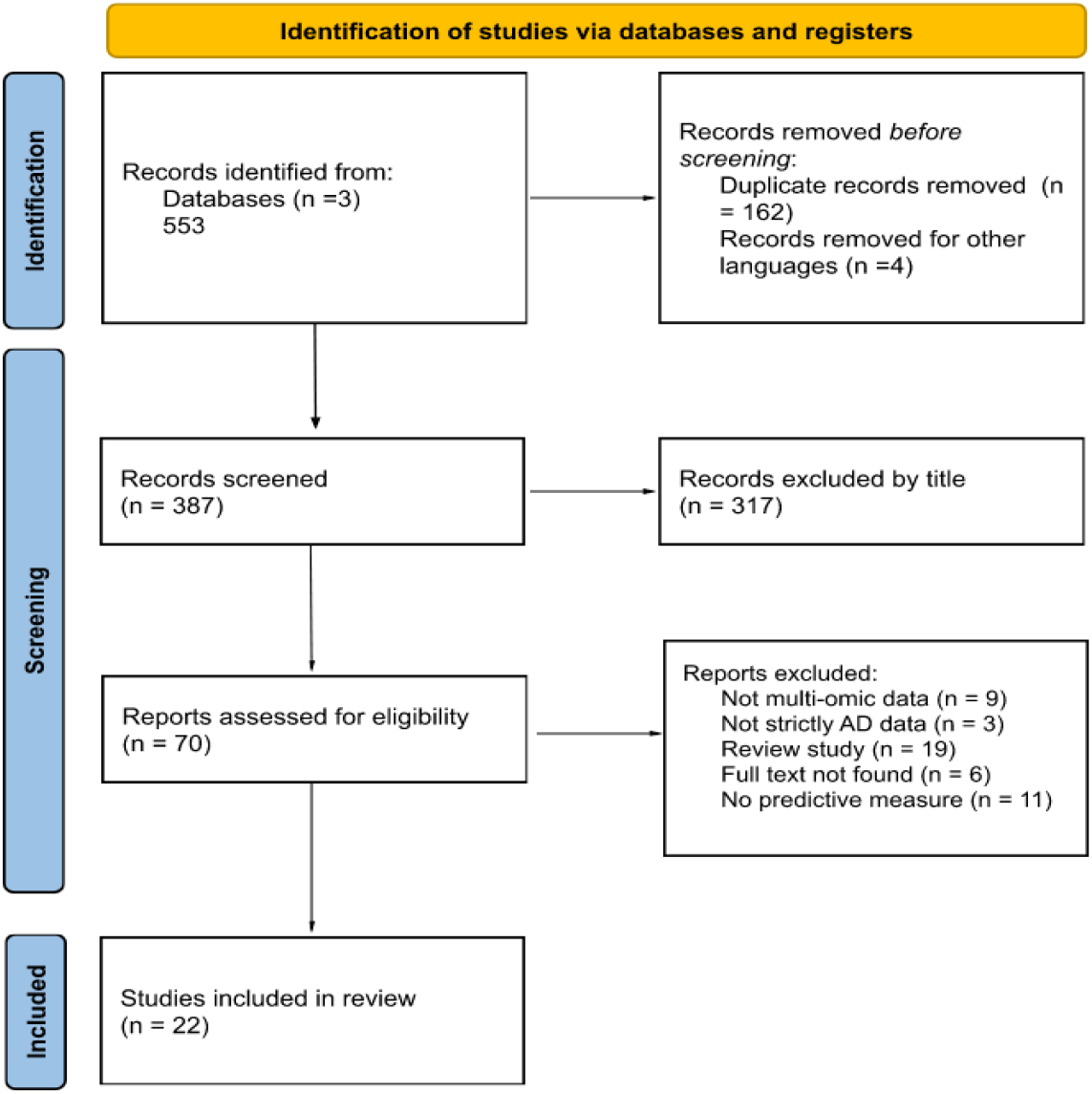
Study Selection Flow Diagram.

### Excluded Studies

For those studies that were excluded after reviewing the abstracts, the reasons for their exclusion were: They did not use multi-omic data The study did not focus on AD explicitly It was a review study The full text could not be found There was no explicitly stated metric for model prediction performance nor biomarker identified

### Included Study Characteristics

Twenty-two studies were included in the final review. Sixteen of the studies included an analysis including genomics or transcriptomics, which were the most common omic data types used. Second most common was proteomics which was included in 7. Other omic data types were neuroimaging (generally MRI scans), metabolomics, lipidomics, methylomics, cytokinomics, clinical test data, and cerebrospinal fluid data.

Studies varied widely in their approach to predicting AD. Some, such as Yu et al. [17], Shigemizu et al. [20], Gupta et al. [21], and Binder et al. [22], aimed to identify biosignatures, a specific combination of biomarkers, which together would predict biomarkers. Others were more general in their biomarker identification, highlighting hundreds of biomarkers which are associated with AD, such as in Maddalenda et al. [23], Song et al. [24][23], Clark et al. [25], Darst et al.[26] [33], Khullar and Wang [27], and Corce et al. [28]. A third category emerged which did not aim to identify specific biomarkers, but instead focused on creating a model which would predict AD based on a continuous feature representation in a machine learning model, such as Abbas et al. [29] and Venugopalan et al. [30].

Types of models common to these studies are the SVM, RF, and XGBoost. Bespoke models published in these studies include DeepGAMI [31], JDSNMF [32], MSLPL [33] and Venugopalan’s autoencoder and CNN.

The most commonly cited dataset in this survey was the Alzheimer’s Disease Neuroimaging Initiative (ADNI), which is a comprehensive longitudinal dataset based in the US with genomics, images, clinical data, and biospecimens. Another popular dataset was the Religious Orders Study/Memory and Aging Project (ROSMAP), which is another multi-omic longitudinal dataset that includes genomics, transcriptomics, methylomics, proteomics, and metabolomics. Another notable source of data was the Gene Expression Omnibus, which is a database for gene expression profiling and RNA methylation profiling. Two studies used primary collected data in their analysis or validation, Corces et al. and Yu et al.. Beckman et al. [34] was the only study to use mouse models to validate their conclusions in vivo.

### Risk of Bias in Studies

Because of the variety in the approaches used in the studies included in this review, it was difficult to assess bias consistently. The studies did not approach predicting AD in the same way, and therefore emphasize different aspects of their research. However, there were common themes between some studies which caused the reviewers to rate 6 of the 22 papers with a medium or high risk of bias. The only study with a high risk of bias was Abdullah et al. [35], which reported a prediction accuracy of 100% with a dataset of only 47 individuals and no reported means of data validation. The other 5 medium risk of bias studies were the papers of Yu et al., Darst et al., Binder et al., Khullar and Wang, and Francois et al.[36]. Khullar and Wang, Binder, and Darst had unclear sample populations, with no reporting of the percent incidence of AD in the datasets. Other concerns in Francois et al. and Darst et al. were the lack of any data validation and a non-diverse dataset. Datasets which used a non-diverse dataset were considered low risk if they included some form of unbiased data validation. Yu’s study was conducted appropriately but included only 31 total samples in the analysis and therefore is at a medium risk of bias.

**Table 1:**
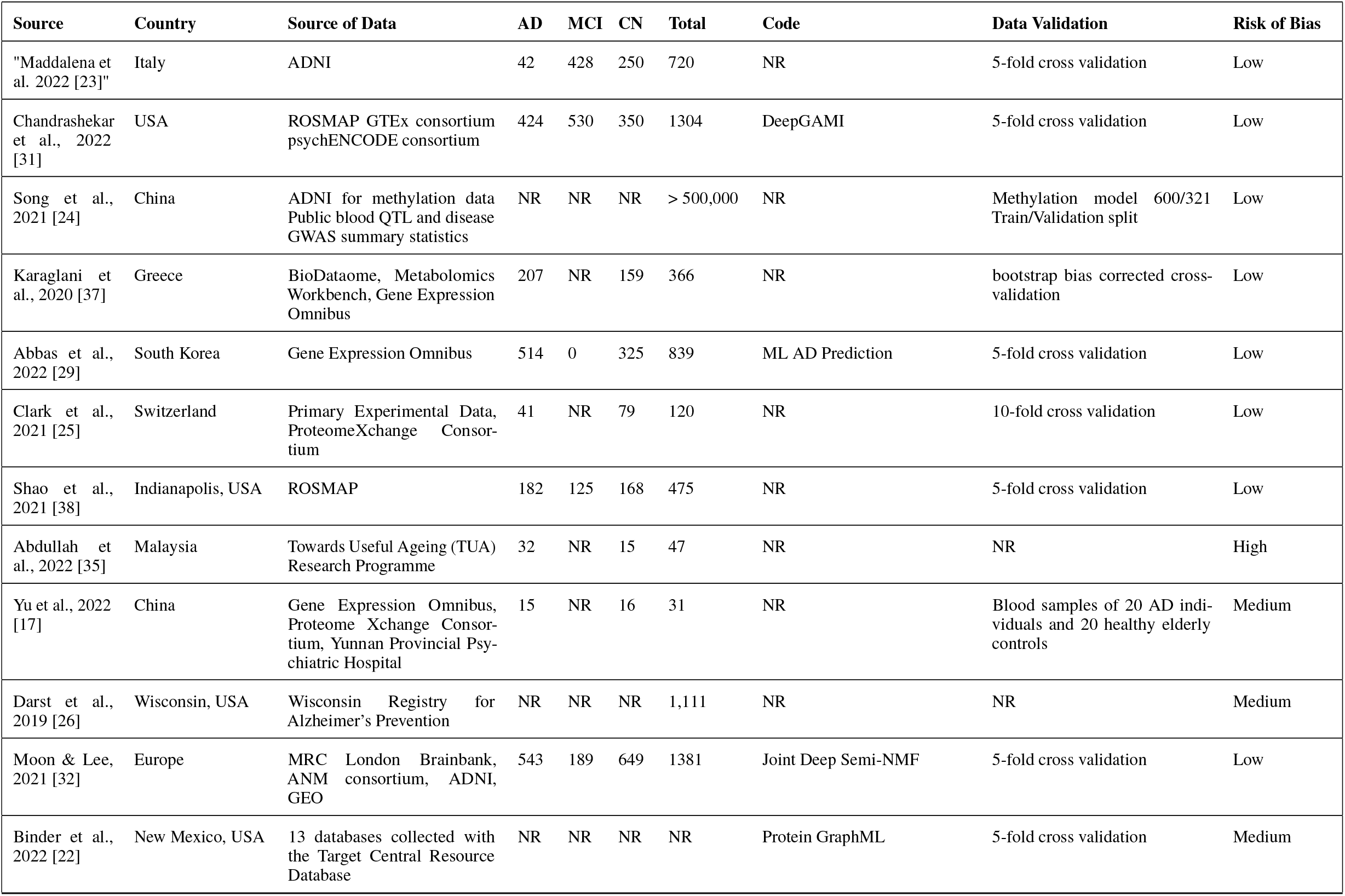
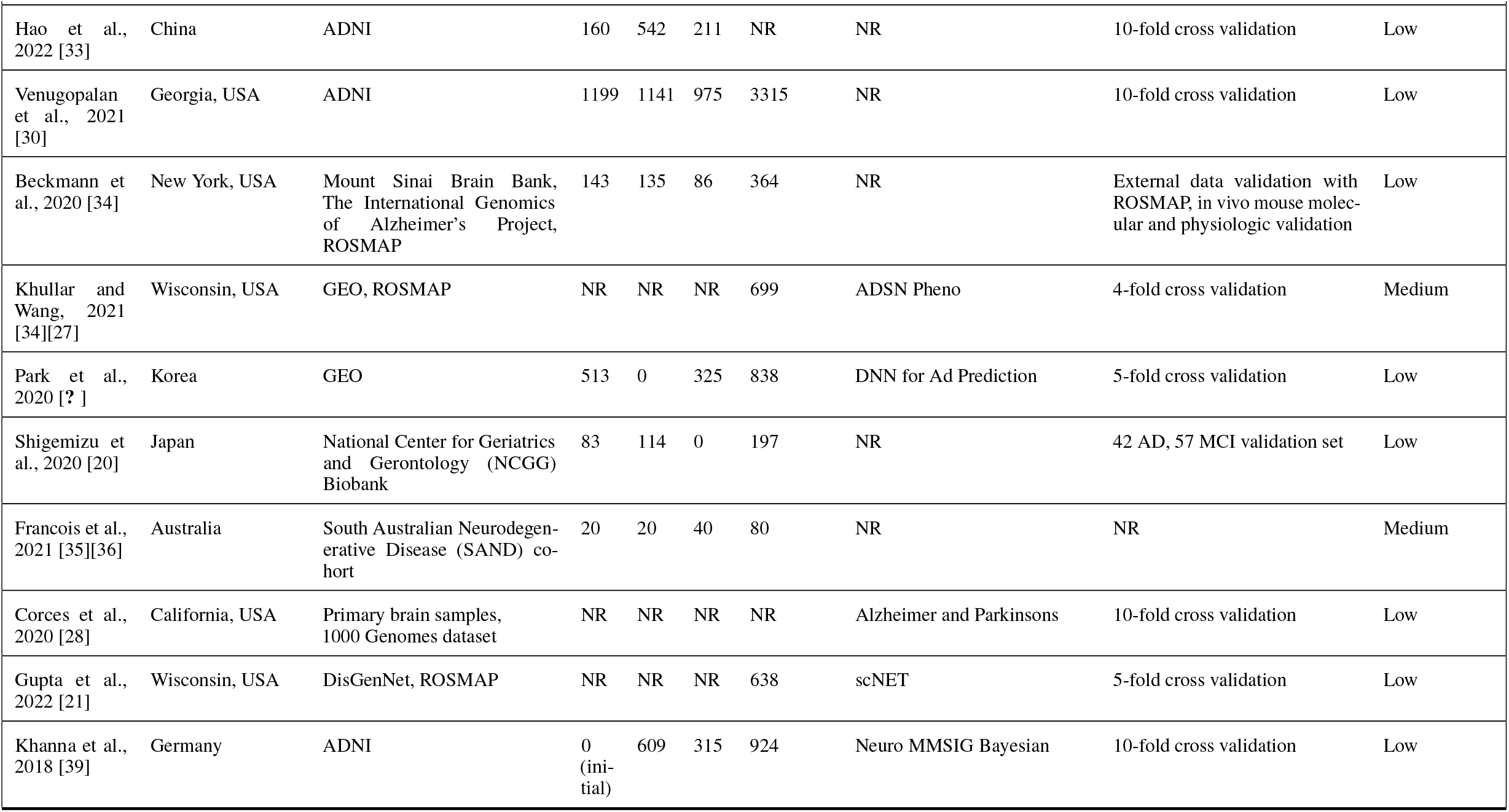
Risk of bias of individual studies

| Source | Country | Source of Data | AD | MCI | CN | Total | Code | Data Validation | Risk of Bias |
| --- | --- | --- | --- | --- | --- | --- | --- | --- | --- |
| "Maddalena et al. 2022 [23]" | Italy | ADNI | 42 | 428 | 250 | 720 | NR | 5-fold cross validation | Low |
| Chandrashekar et al., 2022 [31] | USA | ROSMAP GTEx consortium<br>psychENCODE consortium | 424 | 530 | 350 | 1304 | DeepGAMI | 5-fold cross validation | Low |
| Song et al., 2021 [24] | China | ADNI for methylation data<br>Public blood QTL and disease<br>GWAS summary statistics | NR | NR | NR | > 500,000 | NR | Methylation model 600/321<br>Train/Validation split | Low |
| Karaglani et al., 2020 [37] | Greece | BioDataome, Metabolomics<br>Workbench, Gene Expression<br>Omnibus | 207 | NR | 159 | 366 | NR | bootstrap bias corrected cross-<br>validation | Low |
| Abbas et al., 2022 [29] | South Korea | Gene Expression Omnibus | 514 | 0 | 325 | 839 | ML AD Prediction | 5-fold cross validation | Low |
| Clark et al., 2021 [25] | Switzerland | Primary Experimental Data,<br>ProteomeXchange Consor-<br>tium | 41 | NR | 79 | 120 | NR | 10-fold cross validation | Low |
| Shao et al., 2021 [38] | Indianapolis, USA | ROSMAP | 182 | 125 | 168 | 475 | NR | 5-fold cross validation | Low |
| Abdullah et al., 2022 [35] | Malaysia | Towards Useful Ageing (TUA)<br>Research Programme | 32 | NR | 15 | 47 | NR | NR | High |
| Yu et al., 2022 [17] | China | Gene Expression Omnibus,<br>Proteome Xchange Consor-<br>tium, Yunnan Provincial Psy-<br>chiatric Hospital | 15 | NR | 16 | 31 | NR | Blood samples of 20 AD indi-<br>viduals and 20 healthy elderly<br>controls | Medium |
| Darst et al., 2019 [26] | Wisconsin, USA | Wisconsin Registry for<br>Alzheimer's Prevention | NR | NR | NR | 1,111 | NR | NR | Medium |
| Moon & Lee, 2021 [32] | Europe | MRC London Brainbank,<br>ANM consortium, ADNI,<br>GEO | 543 | 189 | 649 | 1381 | Joint Deep Semi-NMF | 5-fold cross validation | Low |
| Binder et al., 2022 [22] | New Mexico, USA | 13 databases collected with<br>the Target Central Resource<br>Database | NR | NR | NR | NR | Protein GraphML | 5-fold cross validation | Medium |
| Hao et al., 2022 [33] | China | ADNI | 160 | 542 | 211 | NR | NR | 10-fold cross validation | Low |
| Venugopalan et al., 2021 [30] | Georgia, USA | ADNI | 1199 | 1141 | 975 | 3315 | NR | 10-fold cross validation | Low |
| Beckmann et al., 2020 [34] | New York, USA | Mount Sinai Brain Bank, The International Genomics of Alzheimer's Project, ROSMAP | 143 | 135 | 86 | 364 | NR | External data validation with ROSMAP, in vivo mouse molecular and physiologic validation | Low |
| Khullar and Wang, 2021 [34][27] | Wisconsin, USA | GEO, ROSMAP | NR | NR | NR | 699 | ADSN Pheno | 4-fold cross validation | Medium |
| Park et al., 2020 [? ] | Korea | GEO | 513 | 0 | 325 | 838 | DNN for Ad Prediction | 5-fold cross validation | Low |
| Shigemizu et al., 2020 [20] | Japan | National Center for Geriatrics and Gerontology (NCGG) Biobank | 83 | 114 | 0 | 197 | NR | 42 AD, 57 MCI validation set | Low |
| Francois et al., 2021 [35][36] | Australia | South Australian Neurodegenerative Disease (SAND) cohort | 20 | 20 | 40 | 80 | NR | NR | Medium |
| Corces et al., 2020 [28] | California, USA | Primary brain samples, 1000 Genomes dataset | NR | NR | NR | NR | Alzheimer and Parkinsons | 10-fold cross validation | Low |
| Gupta et al., 2022 [21] | Wisconsin, USA | DisGenNet, ROSMAP | NR | NR | NR | 638 | scNET | 5-fold cross validation | Low |
| Khanna et al., 2018 [39] | Germany | ADNI | 0 (initial) | 609 | 315 | 924 | Neuro MMSIG Bayesian | 10-fold cross validation | Low |

### Results of Individual Studies

The studies in this survey took different approaches to how they approach the problem of AD prediction, and therefore did not measure their success with the same metrics. Generally, some of the studies focused more on the biomedical reporting, while others focused on a statistical or machine learning approach, but they all shared some aspects of each. Of the 22 studies, 15 reported an AUC metric for their model. It is not fair to directly compare AUC between these studies because each of the reported metrics used different datasets with different progressions of AD and MCI. Some studies such as Clark and Hao focused on differentiating between MCI and AD and were therefore more difficult than those differentiating AD and CN, which is more obvious to separate. Regardless, 12 of the 24 studies reported a biomarker, collection of biomarkers, or feature representation that can predict AD with 80% AUC or better.

The AUC scores achieving over 90%, baring Abdullah’s score of 100%, were from Karaglani et al. (98%), Clark et al. (98%), Maddalenda et al. (95.5%), Hao et al. (95%), Binder et al. (93%), Kuller and Wang (92.5%), Chandrashekar et al. (92%), Abbas et al. (90%). All these scores were achieved in differentiating healthy patients from those with symptomatic AD, not MCI; scores delineating MCI from AD were significantly poorer in all studies. Karaglani et al. was most successful in using only miRNA with no other multi-omic data using the Just Add Data Bio platform. Clark et al. identified a successful collection of protein, metabolite and lipid biomarkers with their Multi-Omic Factor Analysis. Maddalenda et al. found their best results extracting brain image features with a CNN and combining it with gene expression features in an SVM. Hao’s approach was similar to Maddalenda et al. in that it studies multimodal imaging genetics, but also included the development of a self-paced locality preserving learning model. Binder et al. identified 5 previously unsuspecting genes using XGBoost with combined genomic, transcriptomic and proteomic data. Kuller and Wang found 121 AD associated gene co-expression modules using a weighted correlation network analysis. Chandrashekar developed an interpretable deep learning model which uses prior biological knowledge to identify 45 genes, SNPs and TF associated with AD. Finally, Abbas et al. combined methylomics and transcriptomics into a continuous feature space using an autoencoder and makes predictions using XGBoost.

SVM and XGBoost were repeatedly high scoring models, with Maddalena’s SVM achieving .955 AUC using genomics and neuroimaging and Abbas et al. achieving .9 AUC with methylomic and transcriptomic data in XGBoost. KNNs and PCA were frequently used as well for preprocessing data into clusters and simplifying the analysis, as in Shao, Yu and Darst. Three studies opted to produce their own machine learning technique to improve performance over published methods. Chandrashekar et al. produced DeepGAMI, Moon and Lee produced Deep Semi-non-negative Matrix Factorization (JDSNMF), and Hao et al. produced Multi-Modal Selfpaced Locality Preserving Learning (MSLPL). Only Hao et al. did not publish their model open source, while Chandrashekar et al. and Moon and Lee did.

The 7 studies which did not report an AUC did report validated biomarkers that were associated with AD, however the degree to which they are associated is vaguer. It is important to note that causality was not investigated in any study besides Beckman et al., who’s research heavily implies that the gene VGF is a causal player in the development of AD.

Not all studies integrated the multi-omic data types together for their analysis, and the only studies which reported the differences in prediction performance between the individual data types was Karaglani et al., which has great success (.98 AUC) in predicting AD using miRNA, and Venugopalan et al. which surprisingly had better performance using only imaging data and not integrating EHR and SNP data.

Only Shigemizu et al. and Khanna et al. performed a time-based analysis showing the differences in progression of AD in high-risk and low-risk individuals, which is indispensable for investigating a disease that is often hard to differentiate between MCI. Khanna et al. used a gradient boosting machine over 96 months of ADNI data, which saw 4.4% of CN patients and 39% of MCI patients develop AD, with a validation c-index .86. Shigemizu et al. looked at genomics and transcriptomics data of MCI to AD conversion patients over 83 months an achieved a c-index of .702 with a logistic regression model.

**Table 2.** Results of individual studies. Sample sizes are organized by AD progression stage, with sample sizes from different omics datasets totaled together for simplicity. Note, the sample sizes are not necessarily the number of patients, as some studies use partially sample matched data. Reported predictive metrics are either from an external test set or validation set. Best performing methods are reported in the table with the method in parentheses. Model prediction performance in different stages of disease progression is also noted in parentheses.

| Source | Data Types | Biomarkers | Model(s) | AUC | Accuracy | Sensitivity | Specificity |
| --- | --- | --- | --- | --- | --- | --- | --- |
| Maddalena et al. 2022 [23]" | Transcriptomics, Neuroimaging | 300 genes and 385 regional features of MRI images | Support Vector Machine (SVM) | 0.955 | 0.95 | 0.722 | 0.983 |
| Chandrashekar et al., 2022 [31] | Genomics, Transcriptomics | 45 genes, SNPs and TF | DeepGAMI | .73 (CI COGDX), .92 (late stage BRAAK) | NR | NR | NR |
| Song et al., 2021 [24] | Genomics, Transcriptomic, Proteomics, Methylomics, Cytokinomics, and Metabolomics | 524 RNA, 807 methylation sites, 29 proteins, 7 cytokines, and 19 metabolites | Two-Sample Mendelian Randomization | 0.73 | 0.7 | 0.73 | 0.64 |
| Karaglanı et al., 2020 [37] | Genomics, Transcriptomic, Proteomics | 3 miRNA predictors; 25 mRNA predictors; 7 protein predictors | SVM, Classification Random Forest, Ridge Logistic Regression | .98 (miRNA), .85 (mRNA), .92 (proteins) | NR | NR | NR |
| Abbas et al., 2022 [29] | Methylomics, Transcriptomics | Continuous features, no biomarkers to report | SVM, RF, NB, XG-Boost, AE | 0.9 (XGBoost), .9 (RF) | 0.92 (XG-Boost), .91 (RF) | NR | NR |
| Clark et al., 2021 [25] | CSF, Proteomics, Metabolomics, and Lipidomics | 26 proteins, 9 metabolites, and 6 lipids | Multi-Omics Factor Analysis | 0.932 (MCI), .98 (AD) | NR | .79 (MCI), 0.86 (AD) | .88 (MCI), 0.97 (AD) |
| Shao et al., 2021 [38] | Genomics, Transcriptomics, Methylomics | 9 DNA methylation features, 17 RNA-seq features and 5 miRNA-seq features | SVM, KNN | 0.86 | 0.82 | 0.84 | 0.8 |
| Abdullah et al., 2022 [35] | Transcriptomics, Metabolomics, Cytokinomics | 7 metabolites, 2 transcripts, and 9 cytokines. | "Univariable Logistic Regression; Multivariable Logistic Regression" | 1 | 1 | 1 | 1 |
| Yu et al., 2022 [17] | Proteomics, Transcriptomics | 3 protein biomarkers | PCA and KNN into a DEG-associated protein interaction analysis | NR | NR | NR | NR |
| Darst et al., 2019 [26] | Genomics, Metabolomics, CSF, Clinical Data | 171 metabolite-gene relationships, 833 metabolite-AD risk factor relationships | PCA; Spearman ranking of pairwise correlations | NR | NR | NR | NR |
| Moon & Lee, 2021 [32] | Genomics, Transcriptomics, Methylomics, Neuroimaging | 15 genes | Deep Semi-negative Matrix Factorization (JDSNMF) | .80 (AD/NL), .77 (MCI/NL) | NR | NR | NR |
| Binder et al., 2022 [22] | Genomic, Transcriptomics, Proteomics | 2 dark genes, 2 immune system genes, and an MTOR-APP regulation gene | XGBoost, MPXgb(AD) | 0.93 | NR | 0.87 | 0.8 |
| Hao et al., 2022 [33] | Genomics, Neuroimaging | 10 SNPs and 10 ROIs | Multi-Modal Selfpaced Locality Preserving Learning (MSLPL) | .95 (AD/NC)<br>.75 (LMCI/AD),<br>.71(EMCI/AD) | .95 (AD/NC), .82 (LMCI/AD),<br>.77 (EMCI/AD) | .96 (AD/NC), .87 (LMCI/AD),<br>.89 (EMCI/AD) | .94 (AD/NC), .79 (LMCI/AD),<br>.59 (EMCI/AD) |
| Venugopalan et al., 2021 [30] | Genomic, Neuroimaging and Clinical Data | Memory tests, imaging summary scores, brain volumes, and 5 SNPs | AE, 3D CNN | NR | .78 (combined features), .84 (imaging only) | .78 (combined features), .83 (imaging only) | NR |
| Beckmann et al., 2020 [34] | Genomics, Transcriptomics, Proteomics, and Clinical data | Neuropeptide precursor VGF | Bayesian Causal Networks | NR | NR | NR | NR |
| Khullar and Wang, 2021 [27] | Genomics, Transcriptomics, Epigenomics | 121 gene co-expression modules | Weighted correlation network analysis | 0.925 | 0.88 | NR | NR |
| Park et al., 2020 [40] | Transcriptomics, Methylomics | 35 genes | Differentially Expressed Gene, Differentially Methylated Position | 0.823 | NR | NR | NR |
| Shigemizu et al., 2020 [20] | Genomics, Transcriptomics | 4 hub genes | Logistic Regression, Cox Proportional Hazard Model | .702 (c-index) | NR | NR | NR |
| Francois et al., 2021 [36] | Genomics, Proteomics, Metabolomics | 12 metabolites and proteins relating to tyrosine metabolism, pyruvate metabolism, glycolysis, and antigen processing | Soft Independent Modelling of Class Analogy (SIMCA) | NR | NR | NR | NR |
| Corces et al., 2020 [28] | Genomics, Epigenomics, Transcriptomics | 174 SNPs and 48 Loci | Gapped k-mer SVM | NR | NR | NR | NR |
| Gupta et al., 2022 [21] | Genomics, Proteomics, Transcriptomics | Microglia TF network dysregulation | RF | NR | 0.62 | NR | NR |
| Khanna et al., 2018 [39] | Genomics, Clinical Data, Neuroimaging | 25 pathways, SNPs, principal components and clinical baseline variables | Gradient Boosting Machine, Bayesian Network | 0.86 (c-index) | NR | NR | NR |

## Discussion

Recent advances in neuroimaging, brain tissue research and sequencing methods have allowed for better insights into the pathogenesis of AD. It is now clear that AD is a complex, polygenic disease with many attributing factors, and not simply a result of amyloid and tau plaque accumulation [17]. Analyzing patient biological data from multiple systems is a promising route to identifying successful biomarkers because it enables investigating mechanisms of disease. Single-omic studies such as genome-wide association studies (GWAS) are limited because they cannot deduce causation from associations and do not represent the complex network effects of physiological changes.

These studies tended to be divided in two groups, those that are focused on the biomedical biomarker development and those that are focused on the bioinformatic algorithm development. The biomedical papers such as Yu et al., Darst et al., and Francois et al. tended to not be concerned with making predictions and rather interested in biomarkers of AD, while the algorithm development studies such as Chandrashekar et al. and Maddalena et al. were more concerned with making predictions than identifying biomarkers. This speaks to the divide between the computer science and medical domains and calls for better collaboration which will allow AI/ML algorithms to both make predictions and explain their conclusions by identifying which biomarkers are most significant in disease. Computer scientists should consider developing tools for biologists not trained in programming, and biologists should consider developing their skills in programming to enhance their data analysis skills.

Disappointingly, few studies made use of longitudinal data to investigate the progression of AD as a function of time. Only Khanna et al. and Shigemizu et al. developed prognostic models which predicted the rate of development of AD in high risk and low risk patients. More longitudinal data should be used to better understand how the disease progresses prior to the emergence of symptoms so that they can potentially be avoided or delayed, rather than a simple binary classification of AD or non-AD. There is currently little certainty in the prediction of AD prognosis given current clinical methods, but hopefully the development of biomarkers and machine learning methods in this review can help improve this ability.

The common theme frequently repeated throughout these studies is the integrated multi-omics approaches score better in identifying more predictive biomarkers than those consisting of individual feature sets. These biomarkers can not only make better predictions together, but also give researchers clues into the mechanisms of AD pathogenesis, as they paint a more complete picture of the processes involved. However, two of the studies did show good or better prediction accuracy using only single-omic data, Karaglani et al. and Venugopalan et al. Therefore, it should be important to remember that using more data is not always better and researchers should take care to consider performing their analysis in a single-omic and multi-omic approach.

Few new biomarkers were common between the studies in this paper. Age and clinical cognitive test data are often cited in these studies as the most predictive factors of AD. However, age and cognitive tests are only considered important after patients have already become symptomatic. APOE 4 status, which can be measured at any time, is also often cited a most important risk factor and together with age and clinical test data can predict AD with 88% AUC [25]. Additional meta-analysis on this topic should be conducted in order to categorize which new biomarkers are the most effective [18].

Multi-omic data is still relatively limited due to sample availability, hosting constraints, and experimental limitations. There are no ‘complete’ multi-omics datasets, only subsets of omics data which could be collected and stored [41]. Datasets in this review have tended to consist of either small sample-matched multi-omic data collected by the authors or multiple combined datasets consisting of data from different individuals. Improving the availability of sample-matched data will help researchers make connections across domains and uncover deeper understandings of disease.

## Conclusion

This systematic review investigated 22 studies developing methods or biomarkers to predict AD. 16 of the studies were high quality studies with a low risk of bias, while 6 of them suffered issues with unclear sample populations, diversity of data and lack of data validation. Hundreds of new AD biomarkers have been identified across all omic domains since 2018, but further research must be done to highlight those that are most clinically useful. Deep learning algorithms which integrate multi-omic biomarkers and features have proven to be able to predict AD with high accuracy given symptomatic patients, but they have not been able to demonstrate the ability to predict AD reliably prior to the onset of symptoms. In order to prescribe preventative therapies, clinicians must be able to rate patients’ risk of AD while they are still cognitively healthy. Future research should leverage longitudinal multi-omic data to identify biomarkers that can be noninvasively collected and significantly contribute to a comprehensive AD risk factor score.

## Data Availability

All data produced in the present study are available upon reasonable request to the authors

## Abbreviations

AD: Alzheimer’s Disease
MCI: Mild Cognitive Impairment
LMCI: Late Mild Cognitive Impairment
EMCI: Early Mild Cognitive Impairment
CI: Cognitively Impaired
NC: Normal Control
NL: Normal
EHR: Electronic Health Record
miRNA: Micro RNA
SNP: Single Nucleotide Polymorphism
TF: Transcription Factor
SCZ: Schizophrenia
SVM: Support Vector Machine
RF: Random Forest
AE: Autoencoder
CNN: Convolutional Neural Network
AUC: Area Under Curve
PCA: Principal Component Analysis
ROSMAP: Religious Orders Study and Memory and Aging Project
ADNI: Alzheimer’s Disease Neuroimaging Initiative

## Others

### REGISTRATION AND PROTOCOL

This review was not registered.

## COMPETING INTERESTS

No potential competing interest was reported by the authors.

## AVAILABILITY OF DATA, CODE, AND OTHER MATERIALS

Assessment materials are available upon request.

## Notes

### Competing Interest Statement

The authors have declared no competing interest.

### Funding Statement

This study did not receive any funding

